# City-wide metagenomic surveillance of food centres reveals location-specific microbial signatures and enrichment of antibiotic resistance genes

**DOI:** 10.1101/2024.07.28.24310840

**Authors:** Jonathan J.Y. Teo, Eliza Xin Pei Ho, Amanda Hui Qi Ng, Shaun Hong Chuen How, Kern Rei Chng, Yiğit Can Ateş, Muhd Tarmidzi Fau’di, Kyaw Thu Aung, Niranjan Nagarajan

## Abstract

The distribution of microorganisms in built environments with high human traffic, such as food centres, can potentially have a significant impact on public health, particularly in the context of increasing worldwide incidence of food and fomite-related outbreaks. In several major Asian cities, public food centres are the main venue for food consumption and yet we lack a baseline understanding of their environmental microbiomes. We conducted city-wide metagenomic surveillance of food-centre microbiomes in Singapore (16 centres, n=240 samples) to provide a detailed map of microbial (bacteria, archaea, fungi, viruses) as well as non-microbial DNA abundances across two timepoints. Food-centre microbiomes were found to be enriched in food-related DNA signatures compared to other environments such as hospitals and offices, with specific food-microbe associations (e.g. Enterobacteriaceae and fish) and food DNA providing a partial explanation for the microbial profiles observed (44% of variation explained). Machine learning analysis identified a small set of microbial species (n=22) that serve as highly accurate (>80%) location-specific signatures for various food centres, some of which persist even after 3 years. Profiling of antibiotic resistance genes (ARGs) and pathogens identified a surprising enrichment of ARGs in food centres relative to other non-healthcare environments (>2.5×), and an order of magnitude enrichment of key pathogenic species (e.g. *Klebsiella pneumoniae*, *Enterobacter spp*) even compared to hospital environments. These results highlight the contribution of diverse biotic and abiotic factors in shaping the unique microbiome profiles of different food-centre environments, and the potential for using metagenomic surveillance to understand the risk for infections and antibiotic resistance gene transmission.

## Introduction

Globally, the WHO estimates that there are more than 600 million food-borne illnesses every year leading to >400,000 deaths^1^. The rising incidence of antimicrobial resistant infections further compounds this challenge with new and emerging pathogens such as Group B *Streptococcus*^2^ being of significant public health concern. In several major Asian cities (e.g. Singapore, Kuala Lumpur and Bangkok), large public food centres (also known as hawker centres) are popular venues for daily food consumption for a significant proportion of the population^3^. Hawkers centers typically represent an improvement in plumbing infrastructure, water supply and sanitation to reduce the incidence of foodborne illnesses^4^. Nevertheless as communal eating spaces, they also have high foot traffic and sharing of tables, plates and utensils, potentially creating an environment conducive to the spread of pathogens. According to a 2020 report by the Singapore Ministry of Health, there were 1,894 and 473 cases of Salmonellosis and Campylobacteriosis in 2019, and >2500 reported food poisoning cases that were epidemiologically linked to food establishments^5^. Despite improved sanitation, food-related gastroenteritis persists, highlighting the potential risk for disease transmission in public eateries. Enhanced surveillance in food centres through microbiological culture-based and genomic approaches thus has the potential to improve our understanding of the distribution of pathogens and antibiotic resistance determinants in the environment, and inform cleaning protocols and other risk mitigation strategies.

A growing body of work has highlighted the use of metagenomics as a surveillance technology for built environments^6^, particularly in areas where there is high human traffic (e.g. subways^7,8^ and offices^9,10^) or where the risk of infection in vulnerable populations is higher (e.g. hospitals^9,11^ and neonatal ICUs^12^). The International MetaSUB consortium, in particular, has pioneered the global collection of metagenomic datasets from cities worldwide over many years to understand the distribution of environmental microbes and the antibiotic resistance genes (ARGs) that they carry^13^. In addition to 16S rRNA sequencing approaches which provide a convenient and cost-effective taxonomic survey of environmental microbiomes, shotgun metagenomics has been particularly popular for such surveys owing to its ability to provide multi-kingdom characterization of samples as well as information on ARGs. Despite this, relatively few studies have characterized the microbiomes of food centres, beyond the use of traditional culture techniques and PCR analysis for select pathogens and ARGs^14–16^. While a few studies have used 16S rRNA sequencing to characterize bacterial genera in kitchens and slaughterhouses^17,18^, our understanding of food-centre microbiomes in relation to other built-environment microbiomes is still limited. In particular, while food centres are not as aggressively cleaned as hospitals^19^, they still have more frequent cleaning schedules than other public locations such as offices and subways. In addition, the use of antibiotics in food production^20^ has the potential to impact the resistome of food centres where large quantities of food are processed and consumed^21^. Finally, food-associated microbes, particularly from fermented foods that are common in Southeast Asian cuisine^22,23^ (e.g. in fish sauce, fermented bean paste and tempeh), as well as from food spoilage in a hot and humid environment^24,25^, could further lend unique microbial community profiles in food centres.

To address this knowledge gap, we conducted a city-wide metagenomic surveillance of food-centre microbiomes in Singapore (16 centres, n=240 samples) to provide a detailed map of microbial (bacteria, archaea, fungi, viruses) as well non-microbial DNA abundances across two timepoints. Notably, our analysis identified enrichment in food-related DNA in food centres compared to other environments, enabling the identification of specific food-microbe associations (e.g. Enterobacteriaceae and fish), as well as evidence that food associations could provide a partial explanation for the microbial profiles observed (44% of variation explained). Using machine learning methods we identified a small set of microbial species (n=22) that can also serve as highly accurate (>80%) location-specific signatures for various food centres, some of which persist even after 3 years. Profiling of antibiotic resistance genes (ARGs) and pathogens identified a surprising enrichment of ARGs in food centres relative to other non-healthcare environments (>2.5×), and an order of magnitude enrichment of key pathogenic species (e.g. *Klebsiella pneumoniae*, *Enterobacter spp*) even compared to hospital environments. These findings highlight the diverse biotic and abiotic contributions that could shape the unique microbiome profiles of food-centre environments, and highlights the utility of food-centre metagenomic surveillance for future studies aimed at reducing the emergence and spread of food and fomite-associated infections and human gut colonization by multi-drug resistant bacteria.

## Results

### City-wide metagenomic surveillance of food centres highlights enrichment of food-related DNA signatures that partially explain microbial profiles

To cover a diversity of food-centre environments in Singapore, we selected a geographically distributed set of sites located in the central, eastern and western parts of Singapore that span the densely populated urban landscape of the city-state (n=16; **Figure 1A**, **Supplementary File 1**). In each site, 10 tables were swabbed based on established MetaSUB protocols^7^ and subjected to deep shotgun metagenomic sequencing (20 million reads on average/sample, **Methods**), providing data for relatively comparable high-touch sites across various food centres (**Figure 1B**). A second set of 80 swabs were collected from these sites 3 years later to assess temporal stability, providing a complete dataset of 226 metagenomes (<6% of samples failed library preparation, **Methods**). The shotgun sequencing data was classified using a custom database of non-redundant genomes from NCBI to enable identification of microbial (e.g. bacteria, archaea, fungi, viruses) as well as non-microbial (e.g. human, plant, animal) reads, and compared to control samples to identify and remove potential signals of laboratory and reagent contamination (**Methods**; **Supplementary File 2**). While the majority of identified taxa were microbial and bacterial as expected (median relative abundance=77%), a substantial proportion of DNA was also classified to non-microbial origins (median relative abundance=12%, **Figure 1C**) including diverse animal and plant orders (**Supplementary Figure 1**). We noted that several of the most abundant orders within the *Animalia* kingdom all had potential food origins including red meat, chicken, duck, oysters and fish (**Supplementary Figure 1A**), while plant orders highlighted potential food sources that are common in local cuisine including peppers, fleshy and leafy vegetables, rice and wheat (**Supplementary Figure 1B**). Interestingly, manual curation of food-related taxa highlighted that corresponding animal and plant orders were strongly enriched for food-related DNA in food-centre metagenomes (>90%, Fisher’s exact p-value<0.0001; **Supplementary Figure 2**, **Methods**). Correspondingly, the total relative abundance of food-related taxa was also significantly enriched in food-centre metagenomes relative to other locations such as hospital sites^9^, office^9^ and outdoor environments surveyed as part of MetaSUB Singapore^13^ (Wilcoxon p-value<0.01; **Supplementary Figure 3A**), with vegetables, fish and meat representing the dominant groups in food-centres (**Supplementary Figure 3B**). These observations highlight the ability to identify potential food-related DNA sources through metagenomic surveillance with particular relevance to food-centre surveillance.

**Figure 1:**
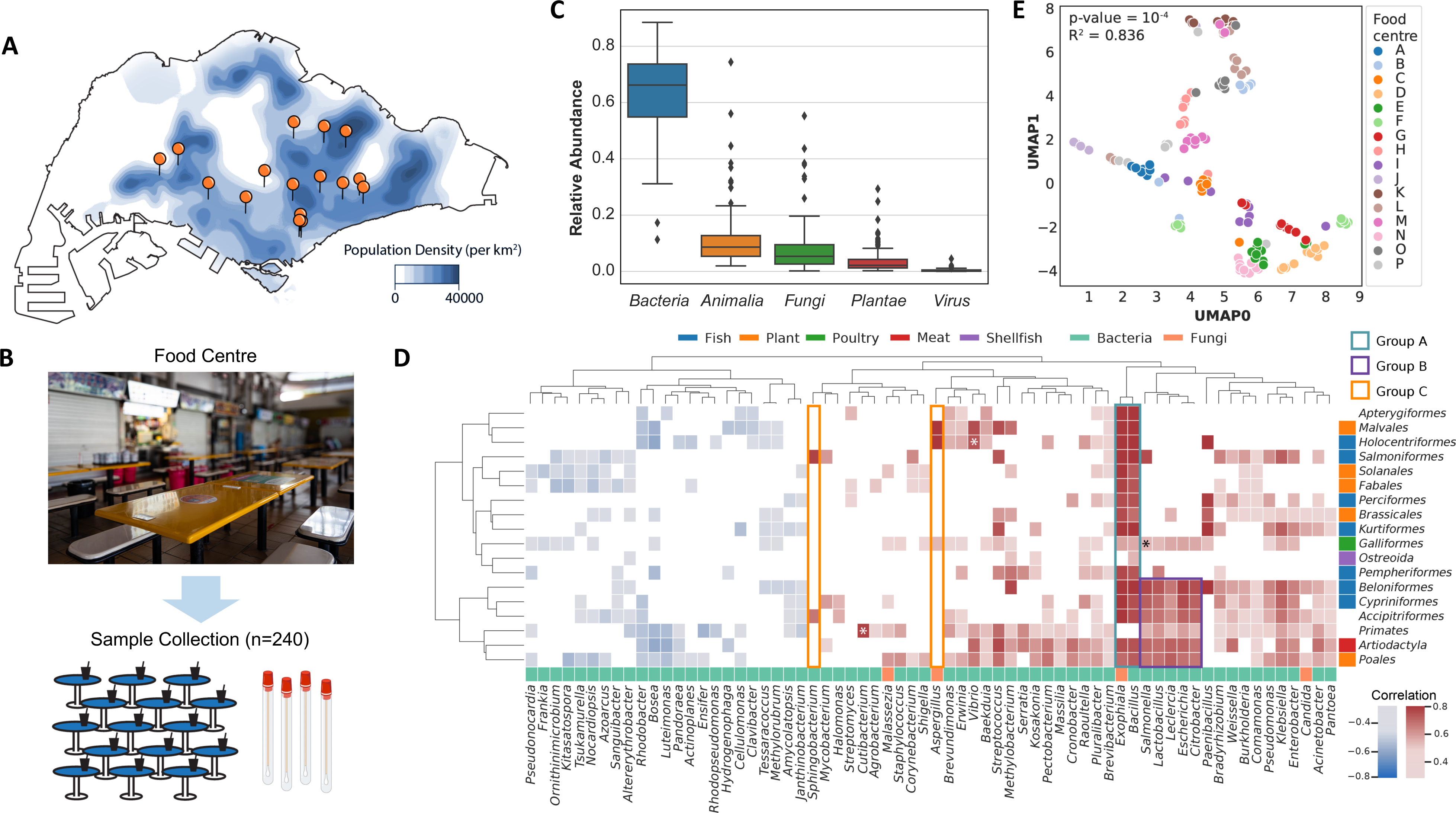
Distribution of microbial and non-microbial taxa in food-centre metagenomes. (A) Map of Singapore showing population density and locations of the 16 food centres that were sampled. (B) Schematic showing a typical food centre setting and sample collection approach for the project. (C) Boxplots showing kingdom-level taxonomic abundances (microbial and non-microbial) based on shotgun metagenomics data (first timepoint). (D) Heatmap of correlation coefficients between microbial genera and non-microbial orders as determined by SparCC (first timepoint; *=key known associations). Only correlations with p-value<0.05 are shown. In addition, only taxa which have at least one correlation value>0.5 are shown. (E) Uniform Manifold Approximation and Projection (UMAP) analysis showing the degree of similarity of taxonomic profiles across food centres based on a Spearman distance metric (first timepoint). PERMANOVA analysis was done based on food-centre labels to obtain the R^2^ and p-value.

We next investigated the potential impact of various food sources on the microbial community using compositionality-aware correlation analysis with SparCC^26^ (**Methods**). As a positive control, common skin commensal species from the *Cutibacterium* genus, were observed to be strongly correlated with the abundance of human DNA, in addition to weaker and non-specific associations of *Vibrio* with fish^27^ and *Salmonella* with poultry^28^ (**Figure 1D**). Some bacterial (e.g. *Bacillus*) and fungal (e.g. *Exophiala*) genera were found to have high correlations with most food-related taxa (Spearman ρ>0.7; Group A, **Figure 1D**; **Supplementary Figure 4**), consistent with their known association with food environments and fermentation^29,30^. Another distinct group of species that includes clinically-relevant opportunistic pathogens from the family Enterobacteriaceae (e.g. *Salmonella*, *Citrobacter*, *Klebsiella*) had strong correlations with a subset of food-related taxa including fish species (e.g. carp), meat and rice (Group B, **Figure 1D**) that are commonly featured in Asian cuisine, a notable association that has not been previously described. Other strong associations were more specific to a small subset of food sources e.g. *Sphingobacterium* with salmon and *Aspergillus* with soldierfish and fruits (Group C; **Figure 1D**), which could indicate a specific source of origin for these organisms. Interestingly, while most of the detected correlations were positive (72%, 300/418), weaker negative correlations were also detected (Spearman ρ<0.5) and were often seen consistently in soil, plant-associated and environmental bacteria (e.g. Frankia, Rhodobacter, Azoarcus). Overall, these results highlight that while some microbes might be associated with food-related DNA abundances in general (**Supplementary Figure 4**), other environmental variables are likely to impact the observed distribution of microbes in food-centre metagenomes as well (percent variation explained by food-related taxa = 44%; **Supplementary Figure 5**).

In terms of microbial taxa, we noted that while bacterial and fungal species from the genus *Klebsiella*, *Enterobacter*, *Acinetobacter*, *Moraxella* and *Candida* were the most commonly seen in food centres metagenomes in this study, the relative proportion of these species varied by location and sampling surface (**Supplementary Figure 6**). For example, while food centres C, E, F and N were marked by high abundance of *Klebsiella* species, food centres A, B, F, K, O exhibited profiles with a higher proportion of *Moraxella* species instead. Overall taxonomic alpha diversities were observed to be higher in food centres relative to other urban environmental metagenomes that we could compare to such as hospital, office and MetaSUB datasets (**Supplementary Figure 7**). Some food centres house market stalls that offer raw ingredients such as meat, vegetables and dairy (**Supplementary File 1**), and while the presence of such stalls was associated with higher microbial diversity (Wilcoxon p-value<0.05; **Supplementary Figure 8A**), the correlation between the number of stalls and Shannon diversity was weak (Pearson ρ=0.17; **Supplementary Figure 8B**). Furthermore, no significant correlation was evident when considering the number of individual food stalls within the surveyed food centres (**Supplementary Figure 8C**), suggesting that other intrinsic factors, such as geography and environment, may play an important role in shaping the higher diversities observed in the metagenomes of these food centres. Intriguingly, microbiomes within the same food centre exhibited greater similarity to each other than to those from different food centres (R^2^=0.873, p-value=10^-4^; **Figure 1E)**. In addition, clustering of microbial taxonomic profiles resulted in clusters that were more similar to location-based groups than clusters based on non-microbial taxonomic profiles (Wilcoxon p-value<10^-4^; **Supplementary Figure 9**; **Methods**). Together, these observations indicate that while variability in food-centre microbiomes might partially reflect food-source variations, food-centre locations and related environmental factors are also likely to play a substantial role in shaping the diversity of microbiota compositions observed.

### Food-centre metagenomes exhibit location-specific microbial signatures that can show long-term persistence

Based on the observation that food-centre metagenomic profiles cluster by location, we next investigated if a machine learning approach could be used to predict locations and identify location-specific metagenomic signatures. As metagenomic data is compositional and high-dimensional in nature, different normalization and feature-selection approaches were explored to construct models (**Figure 2A**; **Methods**). Comparison of performance from different multi-class classification approaches (n=11) highlighted that while high AUC-ROC scores could be achieved (>0.95, 4-fold cross-validation), linear classifiers generally performed better for this task relative to non-linear models (e.g. quadratic discriminant analysis - QDA), and appropriate normalization (ILR: isometric log-ratio transformation, CLR: centered log-ratio transformation) was essential to achieve good performance (**Figure 2B**). Interestingly, logistic regression (LogReg) classifiers exhibited the highest performance across datasets, providing interpretable model weights which are useful for identifying important features for subsequent biological investigations (**Figure 2B**). In addition, microbial features were more predictive of location than non-microbial features, with performance with microbial features alone being similar to the full model, indicating that location-specific metagenomic signatures are primarily microbial in information content (**Figure 2B**).

**Figure 2:**
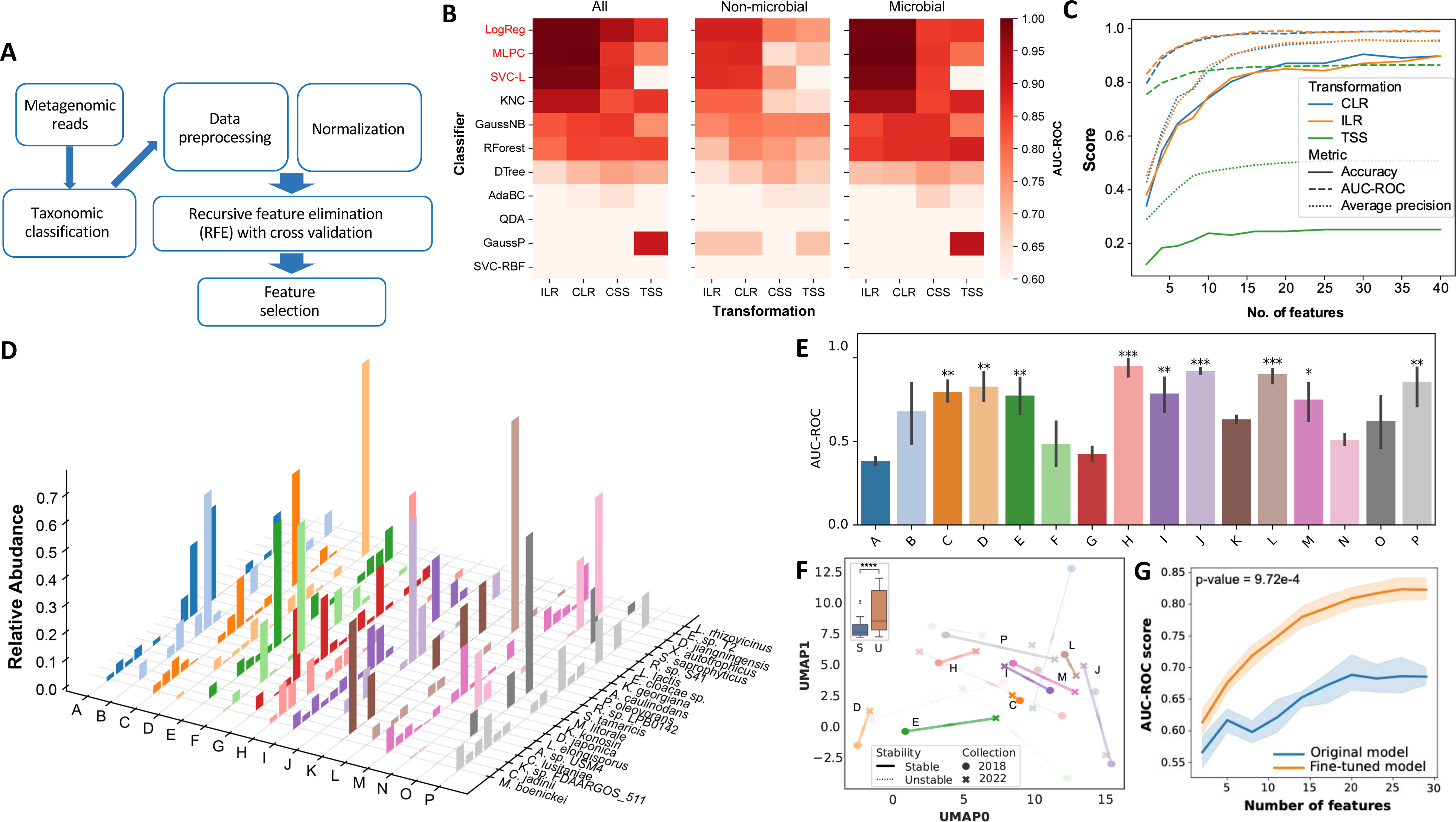
Predicting food-centre locations based on microbial profiles. (A) Schematic of workflow used to assess various normalization techniques and do feature selection. (B) Area-under-curve of the receiver operating characteristic curve (AUC-ROC) for predicting the source of a taxonomic profile using different classifiers (linear models in red text) and normalization techniques. The scores were calculated based on classification of held-out test datasets and 4-fold cross-validation runs. (C) Performance of classifiers with increasing number of features evaluated using 3 scoring metrics - classification accuracy, AUC-ROC score and average precision from the precision-recall curve. Scores are averaged over all food centres. (D) 3D Histogram showing the normalized abundances of 22 microbes that were identified as microbial signatures for various food centres. (E) Boxplots showing the AUC-ROC classification performance across food-centres using a logistic-regression classifier trained with samples from the first collection to classify samples collected 3 years later. Scores are averaged across the classifiers from 4 cross-validation runs. Star notation indicates significant p-values based on the one-sided Wilcoxon rank-sum test (****: p-value<10^-4^, ***: p-value<10^-3^,**: p-value<10^-2^, *: p-value<0.05). (F) UMAP plot of microbial signature profiles from both the first and second collection. Only the centroids from each food-centre cluster are shown. Lines connect the centroids of each food-centre at both timepoints and their opacities increase with shorter distances to indicate higher similarity based on the Spearman distance metric. Inset shows UMAP distances between timepoints for all pairs of tables in food-centres which have stable (S) and unstable (U) classification models (Figure 2E). Star notation indicates significant p-values based on the one-sided Wilcoxon rank-sum test (****: p-value<10^-4^). (G) Comparison of classification performance between classifiers with and without fine-tuning. Fine-tuning describes the process where the model is trained with the original dataset plus a small subset of additional data from the second collection (1 out of 5 samples per food-centre). The mean AUC-ROC scores averaged across food-centres and cross-validation runs are shown (line) along with the 95% confidence interval (shading). Wilcoxon rank-sum test was performed on AUC-ROC scores from models utilizing 20 features.

To identify the essential subset of features, logistic-regression classifiers were trained with varying number of features, highlighting that as few as 20 features could be sufficient to achieve optimal performance (AUC-ROC>0.95, Precision>90%, Accuracy>80%; **Figure 2C**), though the use of principal components generally led to lower performance. The most informative features were then selected using recursive feature elimination with cross-validation (RFECV; **Methods**), yielding 28 species on average across cross-validation runs. Taking the union of species across 4 cross-validation runs identified a core set of 22 species that were consistently selected in at least 3 out of 4 runs (**Table 1**). These 22 species collectively exhibited mean combined relative abundance of 4.5% across all samples, indicating that the identified microbial signatures represent non-dominant species in the metagenome that nevertheless have consistent differences across food centres (**Figure 2D**). Of note, several species are known to have environmental associations (e.g. *Azorhizobium caulinodans* and *Dyella japonica* in soil samples^31^), while other species are known to have associations with foods (e.g. *Clavispora lusitaniae* and *Lactobacillus farciminis* in cheese and kimchi respectively^32,33^) and food additives (e.g. *Cyberlindnera jadinii* for its high glutamic acid content^34^), highlighting the diverse potential sources for these species.

**Table 1:** Details for core species that act as location-specific microbial signatures.

| Species | Location | Mean<br>RA<br>(%) | Habitat | Type | Ref. |
| --- | --- | --- | --- | --- | --- |
| *Mycolicibacterium boenickei* | M | 0.04 |  | Gram+ | 60 |
| *Cyberlindnera jadinii* | D, M | 0.95 | Food | Yeast | 34 |
| *Klebsiella* sp.<br>FDAARGOS_511 |  | 0.04 | Gut | Gram- | 6101/03/2024<br>10:07:00 |
| *Clavispora lusitaniae* | N | 0.18 | Food, Gut | Yeast | 62 |
| *Aquitalea* sp. USM4 |  | 0.05 | Aquatic | Gram- | 63 |
| *Lodderomyces elongisporus* | L | 0.07 | Food, Soil,<br>Blood | Yeast | 64 |
| *Dyella japonica* | C, H, M | 0.40 | Soil | Gram- | 65 |
| *Kushneria konosiri* | F, G | 0.05 | Food (Salt) | Gram- | 66 |
| *Mycolicibacterium litorale* | C, M | 0.05 | Soil | Gram+ | 60 |
| *Salinicola tamaricis* | F | 0.08 | Plant | Gram- | 67 |
| *Rhodobacter* sp.<br>LPB0142 | A, J, P | 0.07 | Aquatic | Gram- |  |
| *Pseudomonas oleovorans* | J, O | 0.13 | Aquatic,<br>lipid-rich | Gram- | 68 |
| *Azorhizobium caulinodans* | A, C | 0.11 | Soil, Plant | Gram- | 31,69 |
| *Kluyvera georgiana* | F | 0.34 | Wastewater,<br>Soil | Gram- | 70 |
| *Enterobacter cloacae* | N | 0.16 | Plant, Soil,<br>Humans | Gram- | 71 |
| *Leuconostoc lactis* | I | 0.04 | Food | Gram+ | 72 |
| *Rhizobium* sp. S41 | G | 0.06 |  | Gram- |  |
| *Staphylococcus saprophyticus* | F | 0.09 | Skin | Gram+ | 73 |
| *Xanthobacter autotrophicus* | C, J, P | 0.06 | Wastewater | Gram- | 74 |
| *Dyella jiangningensis* |  | 0.09 | Soil, Rock | Gram- | 75 |
| *Enterobacter* sp. T2 |  | 0.06 | Wastewater | Gram+ | 76 |
| *Luteibacter rhizovicius* | M | 0.04 | Food | Gram- | 77 |

To assess the temporal persistence of food-centre microbial signatures, we applied the classifier model to a set of metagenomes (n=79) obtained from the same food centres more than 3 years later in 2022. Remarkably, the classifier yielded an AUC-ROC score exceeding 0.7 for food centres C, D, E, H, I, J, L, M and P, which is significantly higher than a permutation-based null model (9/15; **Figure 2E**). In addition, ordination analysis revealed that these food centres (stable – S) had less changes to their microbial profiles compared to other food centres (unstable – U), between the two timepoints of sample collection over a period of three years (**Figure 2F**), further underscoring the persistent nature of microbial signatures in specific food-centres. Next, we fine-tuned the classifiers with a single sample from each food-centre from the second collection (20% of samples) and found a substantial rise in AUC-ROC scores, surpassing an average value of 0.8 (**Figure 2G)**. These results suggest that location-specific microbial signatures in select food centres remain stable over extended periods of time, and in cases where the microbiome has shifted over time, the classifier can be fine-tuned with a small set of new data to improve classification performance.

Generalizing beyond location-specific signatures, we explored if the microbial profiles carried geographical information. Testing for a distance-decay relationship^35^ between microbial profiles (Bray-Curtis dissimilarity) and different food-centre locations in Singapore (Euclidean distance), we observed a weak but statistically significant correlation (ρ=0.082, Mantel test p-value<0.008). Training a classifier based on geographical cluster labels (**Supplementary Figure 10**) achieved slightly lower classification accuracy than for location labels (accuracy= 80%, AUC-ROC=0.9), while using more features (**Supplementary Figure 11A**). Among the features, 8 species were found in at least 75% of the cross-validation runs (**Supplementary Figure 10B**), identifying 3 species that were not identified by the location-based classifier, including a newly isolated species from air samples in Singapore (*Brachybacterium sp. SGAir0954*; **Supplementary Figure 11C**). These results highlight the potential for environmental microbes in food centres to serve as forensic signatures of location and geography, as well as providing an important baseline for identification of potential pathogens and their resistance determinants.

### Enrichment of antibiotic resistance genes and pathogenic species in food-centre microbiomes

To assess the potential role that food-centre microbiomes may play in harbouring antimicrobial resistance determinants and facilitating their onward transmission, we profiled the relative abundance of antibiotic resistance genes (ARGs) in food-centre metagenomes (**Methods**). Food-centre metagenomes harboured relatively higher ARG abundance compared to other environmental metagenomes including office environments and outdoor environments profiled as part of a MetaSUB consortium study (>2.5×, Wilcoxon p-value<10^-4^; **Figure 3A**). Hospital environments were the sites that exhibited highest median relative abundance of antibiotic resistance genes (**Figure 3A**), and this was true across different resistance gene classes, except for colistin resistance which was most enriched in food centres (>1.5×, Wilcoxon p-value<10^-4^; **Figure 3B**). This is noteworthy, as colistin is often an antibiotic of last resort and its indiscriminate use in animal husbandry has been seen as a risk factor for transmission of resistance to human pathogens^36^. Interestingly, for a few other classes of antibiotic resistance (e.g. aminoglycosides, beta-lactamases and Fosfomycin), metagenomes of food-centres, a built environment where high human traffic is typically expected, resembled hospital environments in having higher median abundances than office and MetaSUB sites (**Figure 3B**, **Supplementary Figure 12**), highlighting the importance of food-centre microbiomes as sites for one-health surveillance. While overall levels of beta-lactamase resistance genes were similar, the profiles of specific resistance genes detected were found to belong to 3 major clusters, with cluster 1 being shared between food-centres and hospitals, cluster 2 being unique to food-centres, and cluster 3 being predominantly seen in hospitals (**Figure 3C**). In particular, beta-lactamase genes commonly found in pathogenic species such as *Acinetobacter baumannii*^37^, and *Enterobacter cloacae*^38^ (e.g. bla_ADC_, bla_CMH_, bla_ACT_, bla_MIR_) were relatively more abundant in food centres than in hospitals (Cluster 2; **Figure 3C**), suggesting that these pathogens may be enriched in food centres. Additionally, Carbapenemase genes (e.g. bla_OXA_) were also detected in both food centres and hospitals, but while several bla_OXA_ variants from hospitals have known associations with plasmids, none of the bla_OXA_ genes detected in food centres were linked to plasmids (**Supplementary Figure 13**), indicating that the risk of plasmid-mediated transmission may be lower in food-centres than in hospitals.

**Figure 3:**
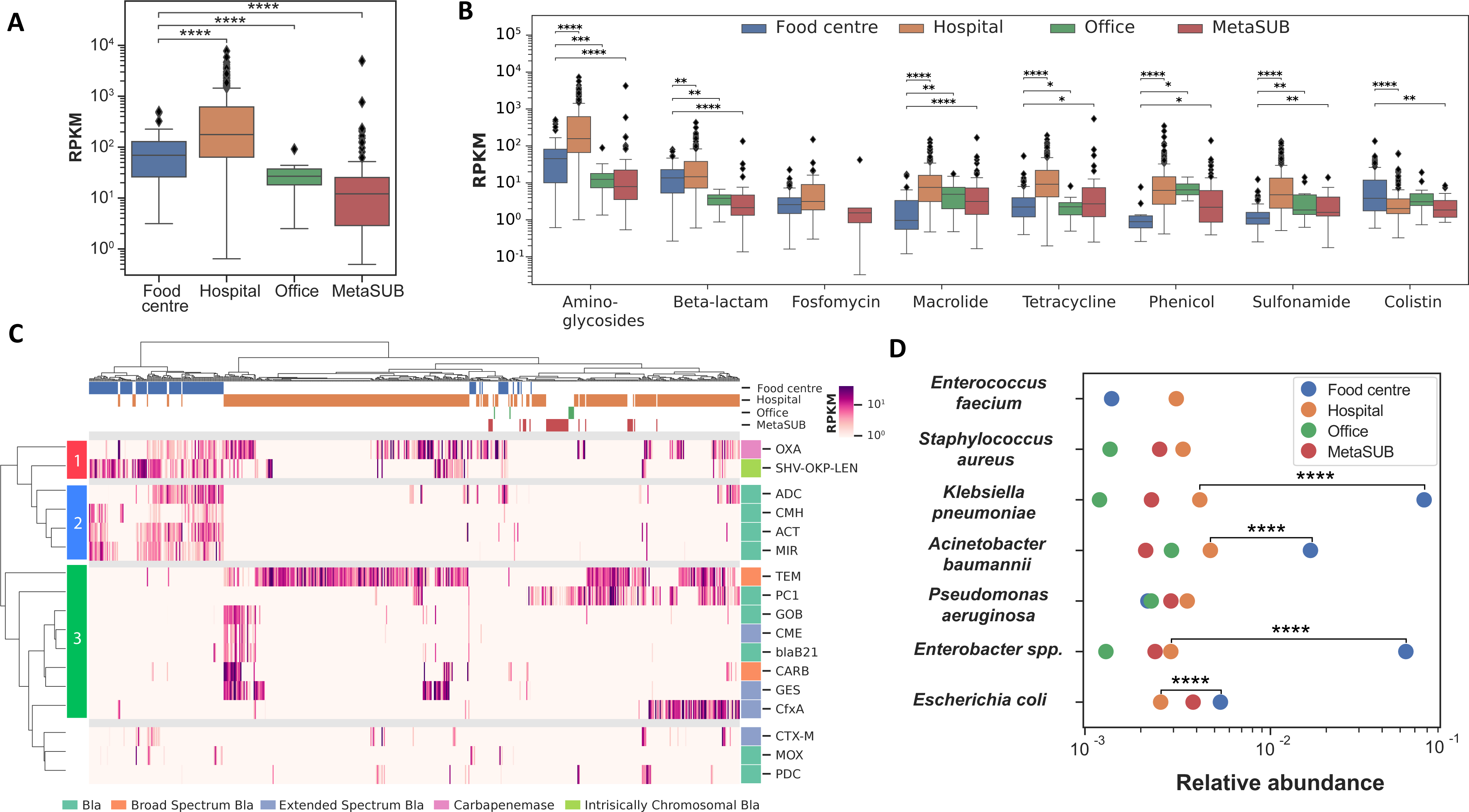
Distribution of antibiotic resistance genes and pathogens across food centres. Boxplots showing (A) antibiotic resistance gene (ARG) abundance (reads per kilobase per million, RPKM) across 4 location classes: food-centres, hospital sites, office and outdoor environments surveyed as part of MetaSUB Singapore (n=221, 429, 23 and 99 respectively; two-sided Wilcoxon rank-sum test, ****: p-value<10^-4^). (B) ARG abundance (RPKM) for various classes of antibiotics across the 4 different locations. Statistically significant differences between food-centres and other locations are shown (two-sided Wilcoxon rank-sum test, ****: p-value<10^-4^, ***: p-value<10^-3^,**: p-value<10^-2^, *: p-value<0.05). (C) Hierarchically-clustered heatmap showing carbapenemase gene abundances (RPKM) in various locations. (D) Strip plots showing the median relative abundance of each ESKAPE pathogen across 4 different location classes (two-sided Wilcoxon rank-sum test, ****: p-value<10^-4^).

We next assessed the abundance of high-priority ESKAPE pathogens (*Enterococcus faecium, Staphylococcus aureus, Klebsiella pneumoniae, Acinetobacter baumannii, Pseudomonas aeruginosa, Enterobacter spp*. and *Escherichia coli*) in food-centres relative to other environments and found that they were surprisingly strongly enriched in food-centres (>3×, Wilcoxon p-value<10^-4^; **Figure 3D**). In particular, gram-negative Enterobacteriaceae such as *K. pneumoniae, A. baumannii, Enterobacter and E. coli* (but not *P. aeruginosa*) were found to be significantly enriched in food-centres than even in hospital environments (Wilcoxon p-value<10^-4^; **Figure 3D**). Overall, these results highlight the utility of metagenomic surveillance in food-centre environments for understanding the distribution of key pathogens and resistance genes, and for further studies into transmission risk.

## Discussion

Despite their social and cultural importance in many major Asian cities, and increasing concerns over new food and fomite-associated pathogens, large-scale microbial genomic surveillance of food centres have not been reported before. Our study helps establish the feasibility of a shotgun metagenomics approach to rapidly conduct city-wide surveillance (estimated sequencing cost <S$10,000), and highlights the utility of this data for tracking microbes and associated genes of concern in the environment. In addition to microbes, our work shows that metagenomics can reflect specific enrichment of food-associated non-microbial taxa in food-centres (**Supplementary Figure 1-3**), though further work is needed to understand how much this reflects food production and consumption patterns. By jointly detecting microbes and food-associated taxa, metagenomics allows us to discover associations between them that may be relevant for tracking the source of food-related pathogens (**Figure 1D**). For e.g., we noted that while some microbes seem to be non-specifically associated to food-related taxa (such as *Bacillus*, *Exophiala*), potentially reflecting a joint seeding pattern or their general ability to grow better with food-derived nutrients, others have more specific associations consistent with prior studies on a food item of interest (e.g. *Vibrio* species in seafood^27^ and *Salmonella* in poultry^28^). Larger-scale analysis may thus help uncover more such associations, especially those that are for rarer microbial taxa and foods that are unique to a few food-centres. Further advances in databases of microbial and non-microbial genomes^39,40^, development of taxonomic classifiers that are more specific and sensitive^41^, long-read sequencing and genome-resolved metagenomics^42,43^ are expected to further aid such studies.

A notable result from this study is that food-associated non-microbial taxa can potentially explain a substantial fraction, but not all, of the microbial variation observed in food-centres (44%; **Supplementary Figure 5**), indicating the importance of food sources in determining food-centre microbiomes. Additionally, it is also clear that there is a strong location-specific signature in metagenomic profiles, which is primarily microbially driven (**Figure 2B**). Similar “forensic” signatures have been reported in other studies as well^44^, but primarily over larger geographic distances (e.g. cities around the world^8,45^). The training of accurate location classifiers (Accuracy>80%) was facilitated by appropriate pre-processing of relative abundance information from metagenomics, enabling highly compact signatures with as few as 20 species to be learnt (**Figure 2C**). Interestingly, these signatures are not defined by abundant taxa (e.g. *Klebsiella*, *Enterobacter* etc; **Supplementary Figure 6**) but by rarer taxa with diverse possible origins (**Table 1**). The higher biomass collection enable by our sampling protocol, as well as the stringency of our ‘kitome’ removal analysis (**Methods**), suggests that these signatures are not likely to be artefactual. In addition, metagenomic data from the second timepoint, more than 3 years later, highlights that some signatures can be remarkably robust over time (**Figure 2E, 2F**). This is despite large socio-environmental changes (COVID-19 pandemic related), as well as unavoidable technical differences (newer DNA extraction protocols; **Methods**) separating the two timepoints. Nevertheless, it highlights how microbes can serve as biomarkers and indicator species for food-centre locations, with further study needed to understand the reasons for a greater shift in these signatures in some sites (**Figure 2F**). The ability to rescue classifier performance with a few additional datasets suggests that these shifts may not lead to a complete replacement of such signatures (**Figure 2G**). Overall, geographical factors appear to provide only a partial explanation for the observed location-specific signatures (**Supplementary Figure 11**), and larger datasets with detailed environmental information could help tease apart the contributions of diverse local factors in shaping food-centre microbiomes.

Food-centre metagenomes were found to be surprisingly enriched for antibiotic resistance genes relative to other outdoor and indoor environments with high human contact (>2.5×; **Figure 3A**), with the not so surprising exception of hospital environments^9^. Of note, food-centres seem to be more similar to hospital environments in the enrichment of various clinically important antibiotic resistance gene classes (e.g. aminoglycosides, beta-lactamases, Fosfomycin; **Figure 3B**). This could in part be due to antibiotic usage in animal husbandry^36^ and agriculture^46^ globally, though the use of antibiotics to promote growth of animals is not allowed in Singapore and so, for example, the enrichment of colistin resistance is unexpected^47^. Other potential explanations include, higher human traffic than other sites, greater microbial biomass thriving on food-based nutrients, and specific environments and interventions (e.g. cleaning protocols) that enrich for bacteria carrying such resistance genes^48^. While the public health ramifications of this enrichment remain to be determined (e.g. chromosomally integrated carbapenemases in food-centre microbiomes may not be as concerning as plasmid-borne genes detected in hospital environments^49^), it is clear that food-centre environments should be an important node for one-health surveillance efforts. This is particularly the case because of the unexpectedly strong enrichment of several ESKAPE pathogens of concern, even relative to hospital environments (>10× for several gram-negative Enterobacteriaceae; **Figure 3D**), and the need to understand if this has an impact on the risk for transmission and infections. Overall, this study highlights the valuable information that metagenomic surveillance can provide about microbes as well as non-microbes in food-centre environments, serving as the basis for further large-scale cross-sectional and interventional studies to understand the diverse factors that shape them, their impact on public health, and the utility of various cleaning and behavioural intervention strategies to reduce infection risk.

## Materials and Methods

### Sample collection

Microbiome sampling was done on food centre table surfaces using Isohelix DNA Buccal Swabs (SK-4S) based on MetaSUB protocols^7^. Samples were collected from 16 different geographically spread-out locations from around Singapore that represent popular food centres in densely populated regions (**Figure 1A**). Sampling was done at two different timepoints (March 2019 and July 2022) within 1 week, where all sites from the same food-centre were sampled on the same day. The second collection was delayed due to lockdowns during the COVID-19 pandemic period. During the first timepoint, 10 different table surfaces were swabbed (total n=160), while 5 tables were swabbed at the second timepoint (total n=80). Swabbing was done with two swabs applied over separate halves of a table, each for a duration of two minutes, and the samples were combined. Swabs were stored in Zymo DNA/RNA Shield™ immediately after collection and the corresponding tubes were subsequently stored at −80°C prior to DNA extraction.

### DNA extraction and sequencing

Extraction of total DNA from swabs was performed via a combination of mechanical and chemical lysis. Briefly, samples were homogenized on the FastPrep Instrument (MP Biomedicals) at 6 ms^−1^ for 40 s, followed by centrifugation (5mins, 13000 rpm) and Proteinase K treatment of supernatant (56°C for 20mins). Purified DNA was obtained using the Maxwell RSC Blood DNA Kit (Promega, AS1400) for the first collection and QIAamp PowerFecal Pro DNA Kit (Qiagen) for the second collection according to manufacturer’s instructions. DNA concentration was quantified on the Qubit dsDNA HS Assay Kit (Life Technologies, Q32854). Samples with low DNA concentration according to the assay were excluded from the study (First collection, n=13; Second collection, n=1). Metagenomic libraries for all samples were prepared with the NEBNext® Ultra™ II FS DNA kit (New England Biolabs, E7805L) according to the manufacturer’s instructions. Paired-end sequencing (2×150bp reads) was performed on the Illumina HiSeq X™ Ten platform. Negative control samples (n=5, blank swabs in DNA/RNA shield) underwent identical DNA extraction and sequencing procedures.

### Taxonomic abundance analysis

Illumina paired-end reads were first trimmed for adapters and filtered for low quality reads using fastp (v0.20.1 -q 25 -p 20)^50^. The remaining reads were used for taxonomic profiling with Kraken2^51^ (v2.0.8) using a custom index (kraken2build –max-db-size 60000) built from the NCBI NT (non-redundant nucleotide) genome database. Taxonomic abundances at the species and genus level were estimated using Bracken^52^ (v2.5).

Abundance profiles were filtered such that species with relative abundance less than 0.01% were excluded to reduce the impact of false-positive calls. To assess the potential impact of reagent and laboratory contamination on taxonomic profiles, DNA quantification was done for negative control libraries (n=5) and found to be a 100-fold smaller than libraries for all collected food-centre samples, suggesting that reagent/laboratory contamination is unlikely to impact the taxonomic profiles for our relatively high-biomass environments (enabled by 2-swab sampling of the entire table). As a conservative measure to reduce the impact of contaminants, species that were present in negative controls were labelled as putative contaminants if their abundance was inversely correlated (pearson correlation >0.3) with DNA concentration in food-centre metagenomes, and correspondingly removed from food-centre microbiome profiles before further analysis. Species from *Animalia* and *Plantae* were manually curated to be food-related if they were known to be common ingredients in local cuisine (**Supplementary File 3**).

### Taxonomic co-occurrence analysis

Taxonomic co-occurrence relationships between microbial and non-microbial species were inferred through the application of the SparCC^26^ method, as implemented in FastSpar v0.0.10^53^ with default parameters. Correlations with absolute coefficients exceeding 0.3 and a statistical significance threshold of p-value<0.05 were retained. The resulting correlations were visually represented using a heatmap, generated with the seaborn clustermap function in Python, employing the Euclidean distance metric.

### Ordination analysis

Ordination analysis was performed using the Uniform Manifold Approximation and Projection algorithm^54^ (UMAP, n_neighbour=10, min_dist=0.1, n_components=2). The Spearman distance metric, which ranges from 0 to 2, was used to measure similarity between subsets of taxonomic profiles. The selection of Spearman distance metric over a more conventional Bray-Curtis dissimilarity metric was aimed at capitalizing on a rank-based methodology to avoid biasing towards highly abundant species. This step is valuable for measuring similarities between profiles containing only a subset of taxa in a microbiome, such as those representing signature species.

### Canonical correspondence analysis

Canonical correspondence analysis (CCA) was performed using the R package vegan^55^ (v2.6.4) to investigate the relationship between microbial composition variations and food-related taxa abundances. In total, 35 food-related orders were examined (**Supplementary File 3**). Percentage of microbial variation explained by food-related taxa was obtained from the ratio of constrained inertia to total inertia. The biplot and model inertia results are shown in **Supplementary Figure 5**.

### Clustering and cluster similarity

Metagenomes were initially clustered based on their microbial abundance profiles or profiles for food-related taxa. These cluster assignments were subsequently compared both against the true location labels of the samples and among themselves using clusim^56^ (v0.4), with higher scores denoting increased similarity. To assess the robustness of clustering, we conducted 10-fold cross-validation to generate multiple similarity scores across the three pairs of comparisons (microbe-food, food-location, microbe-location; **Supplementary Figure 9**). Additionally, a null model was established through random permutations of cluster labels to compute similarity scores and test for statistical significance (one-sided Wilcoxon text).

### Training of machine learning models

Multi-class machine learning classifiers were trained to predict the most likely source of a sample based on its metagenomic profile. To limit the number of features in the training dataset, we restricted the models to the top 200 species based on median abundances. Taxonomic profiles were normalized using various approaches including total sum scaling (TSS), cumulative sum scaling (CSS, metagenomeSeq^57^ v3.18), centered log-ratio (CLR) and isometric log-ratio (ILR) as described by Thomas et al.. A pseudo count of 1 was applied across the dataset to allow for log transformation. Classifiers were trained using 11 different algorithms, including logistic regression, multi-layer perceptron, support vector classifier (linear basis function), K-nearest neighbor, Gaussian naïve Bayes, random forest, decision tree, Adaboost, quadratic discriminant analysis, Gaussian process, and support vector classifiers (radial basis function), as implemented in the python scikit-learn package. Binary classifiers were adapted for multi-label classification using a one-vs-rest approach. The classifiers were first trained on a randomly selected training dataset (n=110) and applied to a withheld test dataset (n=37) to assess classifier performance. This process was repeated 4 times for different splits of the dataset and the mean accuracy and AUC-ROC scores (one-vs-rest, average using ‘macro’) from the test datasets was reported. Subsequent analysis was limited to the logistic regression classifier as it performed the best. To test for statistical significance of classification scores at each food-centre location, a null model was trained from 5 sets of random permutations of the food-centre labels. A one-sided Wilcoxon rank-sum test was performed subsequently to identify locations with significantly higher classification performance.

### Geographical association analysis

Food-centres were grouped together using hierarchical clustering based on Euclidean distances of their geographical location. Mantel’s test (scikit bio v0.1.3) was performed between a distance matrix of Bray-Curtis dissimilarity scores between taxonomic profiles and a second distance matrix of Euclidean distances calculated from geographical coordinates of the food-centres.

### Resistome analysis

Antibiotic resistance gene (ARG) profiles were calculated using SRST2^58^ (v0.1.4; -- min_coverage 100, hits with identity <99% were filtered out) using a curated version of the CARD database^59^ (v3.0.8). Alignment counts to ARGs were normalized by total reads per sample and gene length to calculate RPKM values. Resistome profiles were aggregated based on the ontology grouping of resistance genes in the CARD database, including grouping by gene class and resistance mechanism.

### Data Availability

Sequencing reads are available from the European Nucleotide Archive under project PRJEB73308. Source code and data for reproducing figures are available under MIT license at: https://github.com/CSB5/food_centre_microbiome.

