## Supplementary Figures for "City-wide metagenomic surveillance of food centres reveals location-specific microbial signatures and enrichment of antibiotic resistance genes"

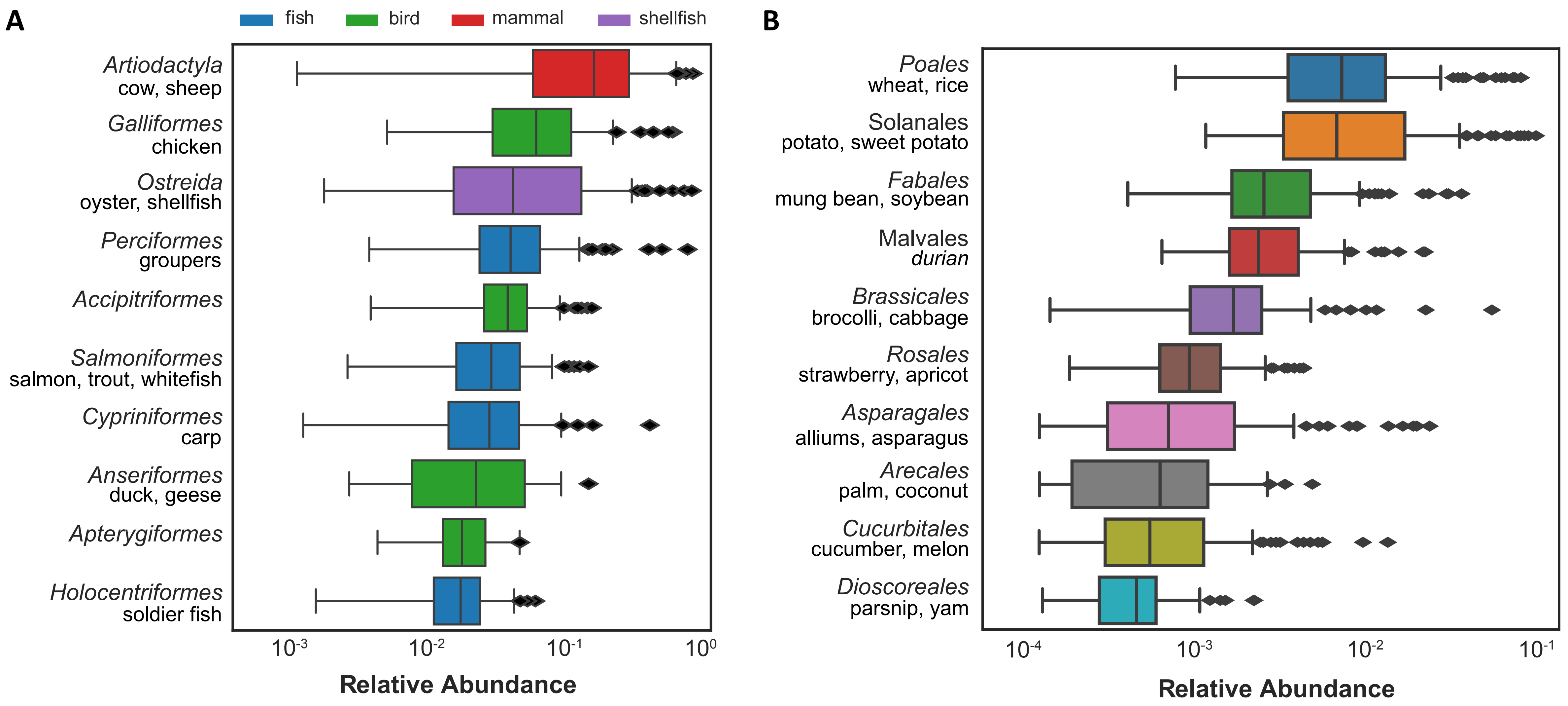


**Supplementary Figure 1:** Boxplots depicting the relative abundances of (A) Animalia and (B) Plantae orders detected in food-centre metagenomes (top 10, sorted by median abundance). Common food-associated species that were detected in each order are listed below their names.

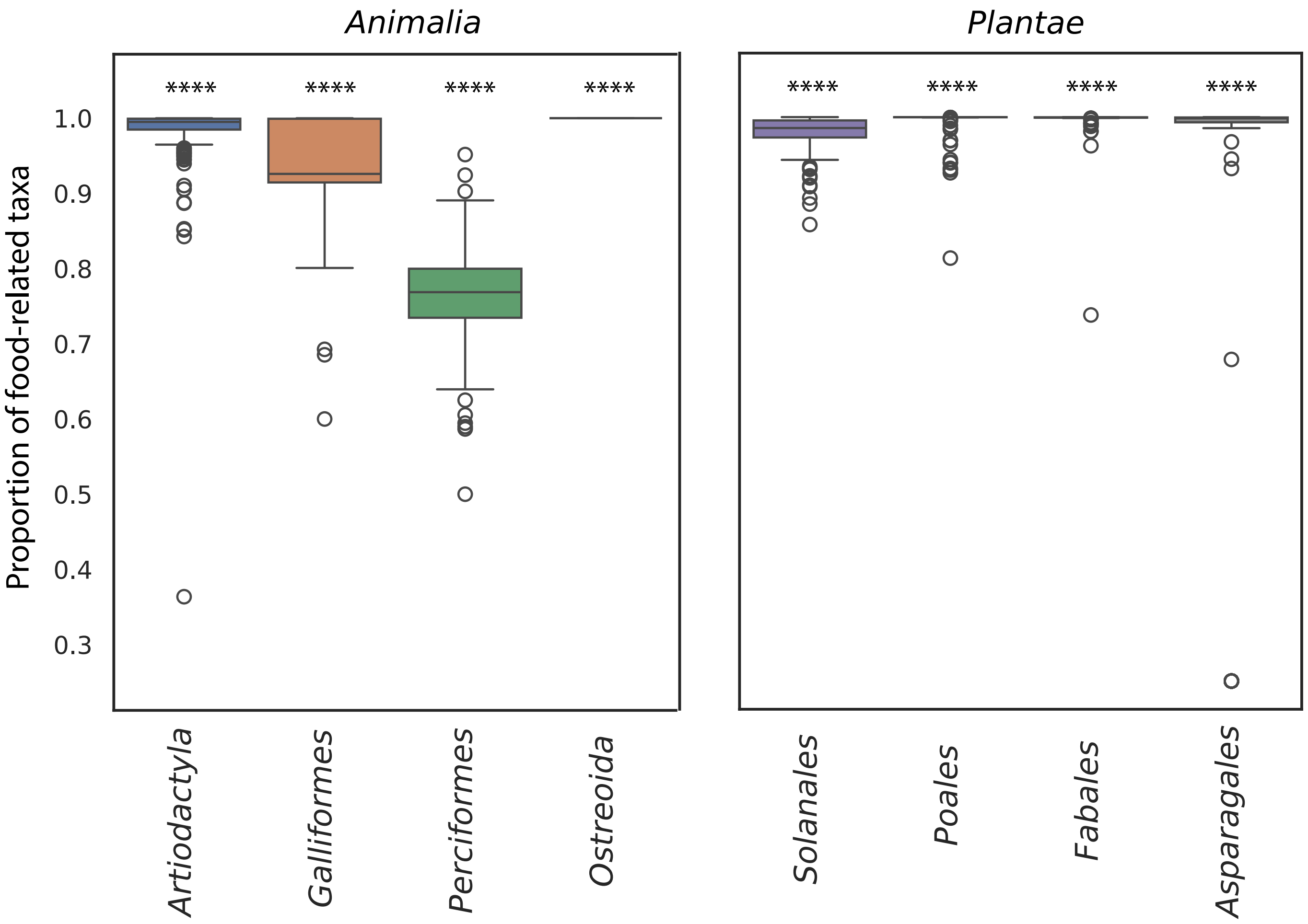


**Supplementary Figure 2:** Proportion of food-related taxa abundances in top four orders from *Animalia* and *Plantae.* Star notation indicates significant p-values based on a Fisher's exact test ("****": p-value<0.0001) compared to a null model where all genera in the order are equally likely to be detected.

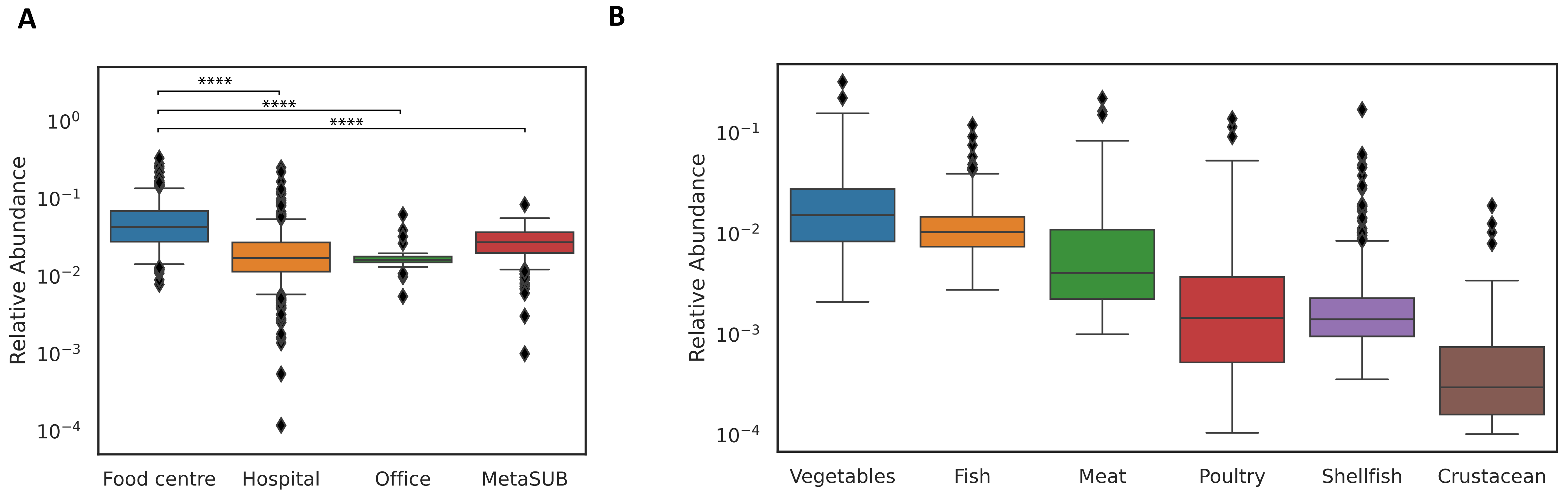


**Supplementary Figure 3:** Relative abundance of food-related taxa, (A) across cohorts, and (B) in food centres that are related to various food sources. Star notation indicates significant p-values based on the two-sided Wilcoxon rank-sum test: "*****": p-value<0.0001.

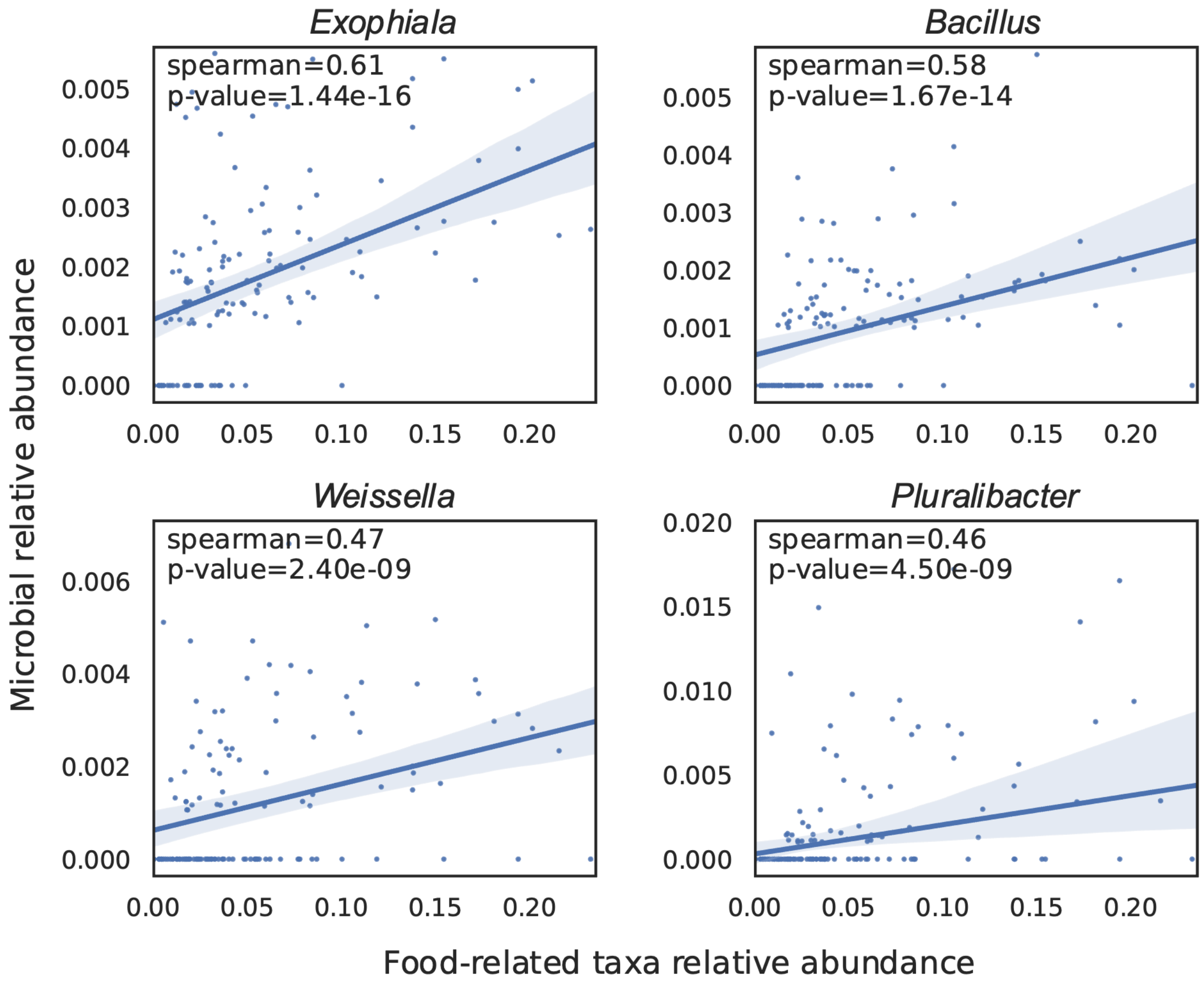


**Supplementary Figure 4:** Top microbial genera that are significantly correlated with total relative abundance of food-related taxa (spearman correlation coefficient >0.4, p-value < 0.05).

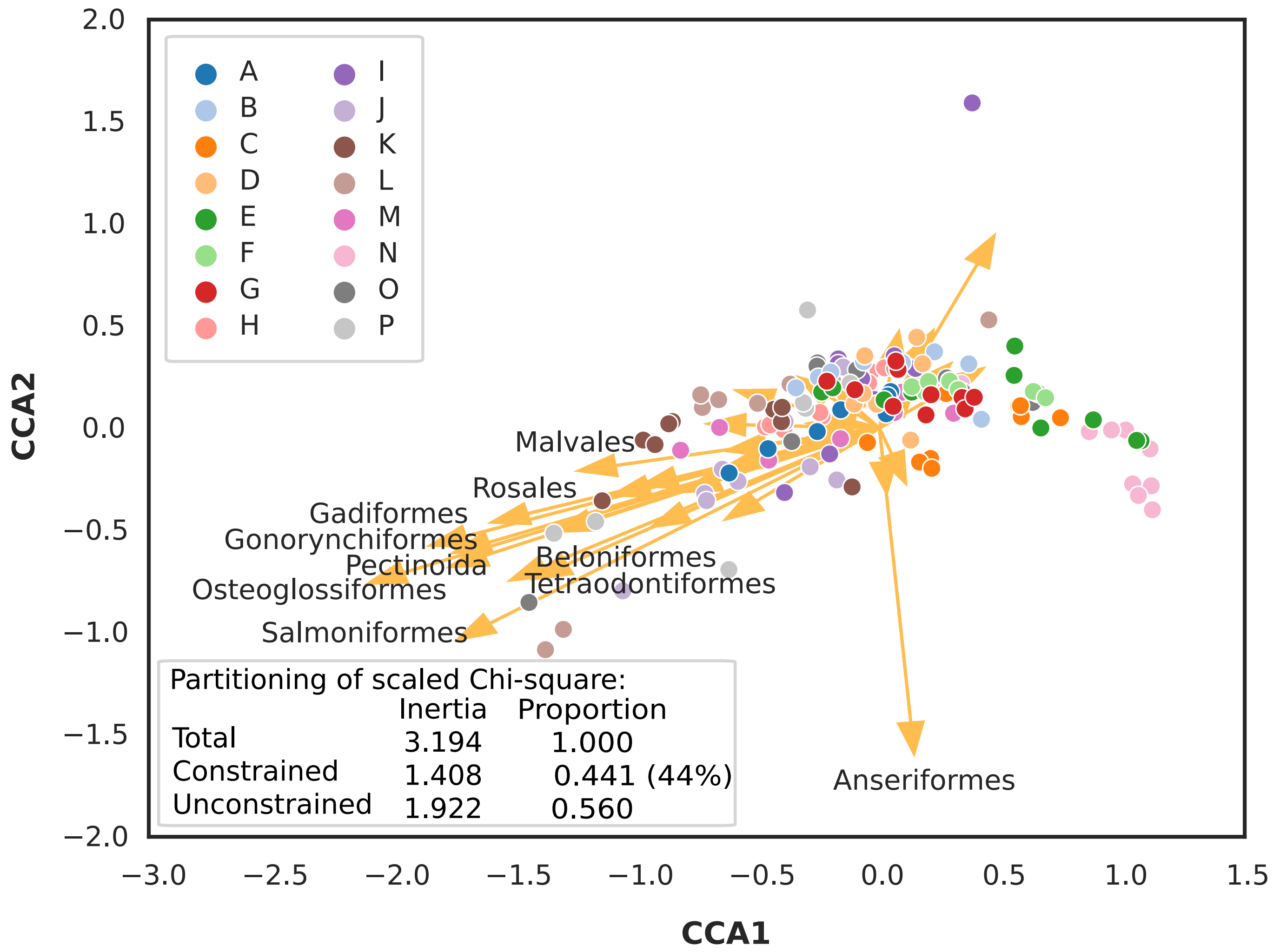


**Supplementary Figure 5:** Biplot from canonical correspondence analysis using food-related orders as environmental variables. Each point represents the microbial community composition from food centres with location indicated by color. Vectors indicate food-related orders used as model constraints, and vectors of magnitude greater than 1 are labelled.

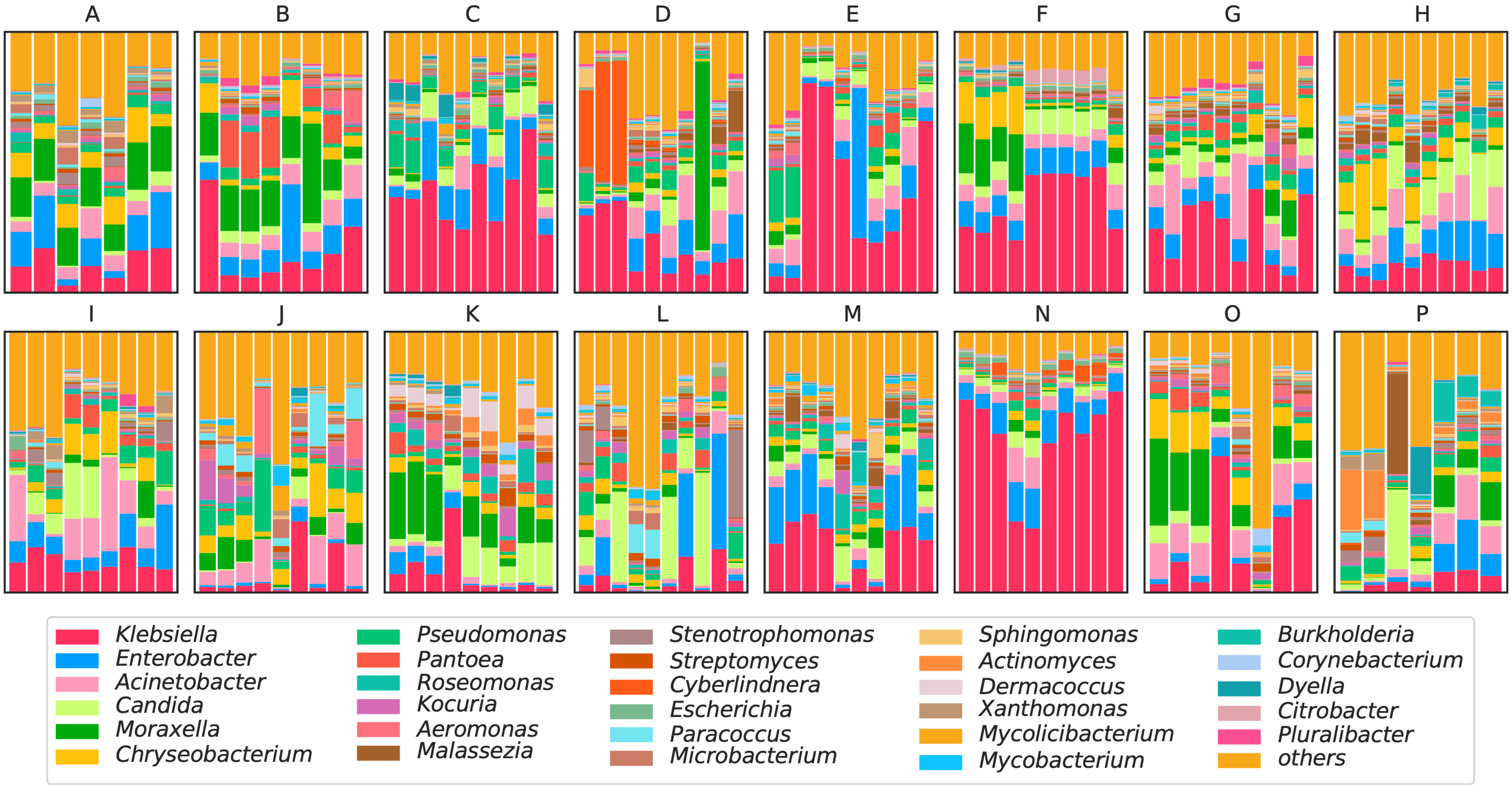


**Supplementary Figure 6:** Stacked barplots of microbial genus-level relative abundances across different sites and food centres (first timepoint). The top 29 genera ranked by geometric mean across samples are shown and the rest are collapsed into 'others'.

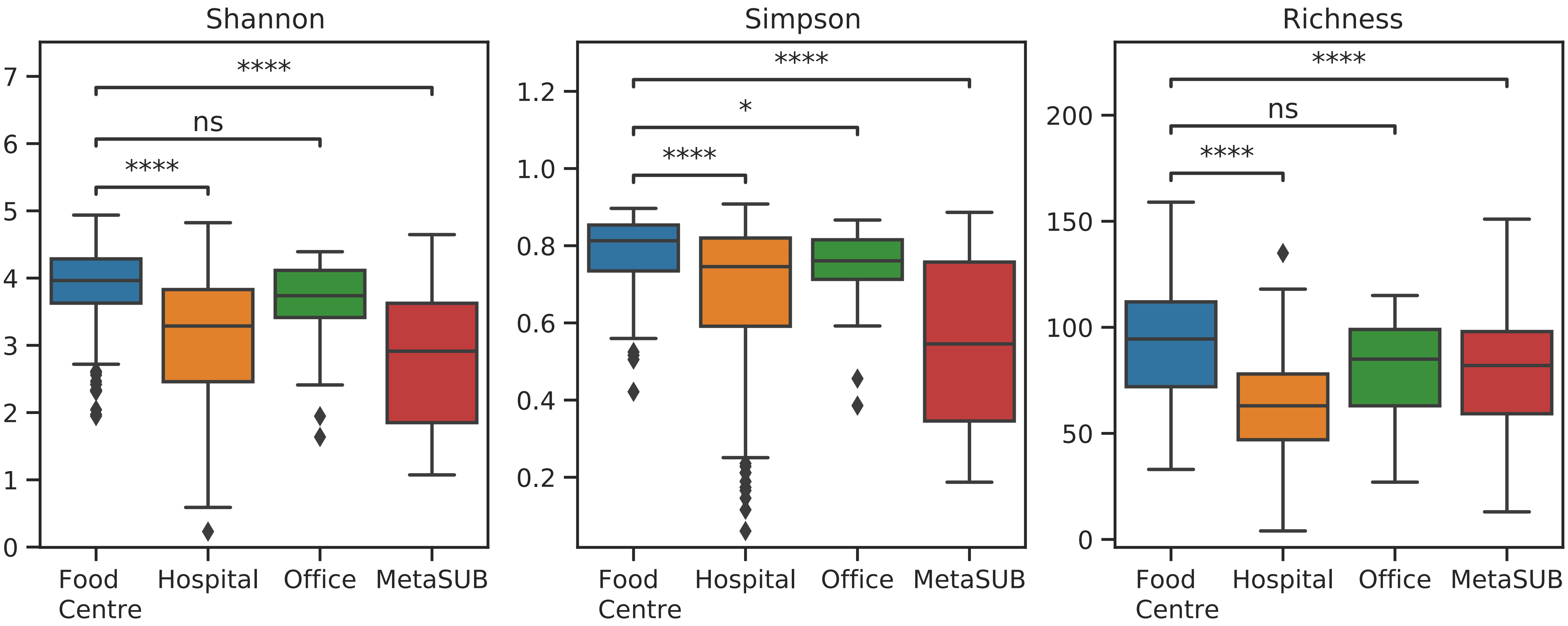


**Supplementary Figure 7:** Boxplots showing alpha diversity metrics for environmental microbiomes in food centre, hospital, office and MetaSUB consortium sites in Singapore. Star notation indicates significant p-values based on the Wilcoxon test: “****”: p-value<0.0001, “*”: p-value<0.05, “ns”: p-value>0.05.

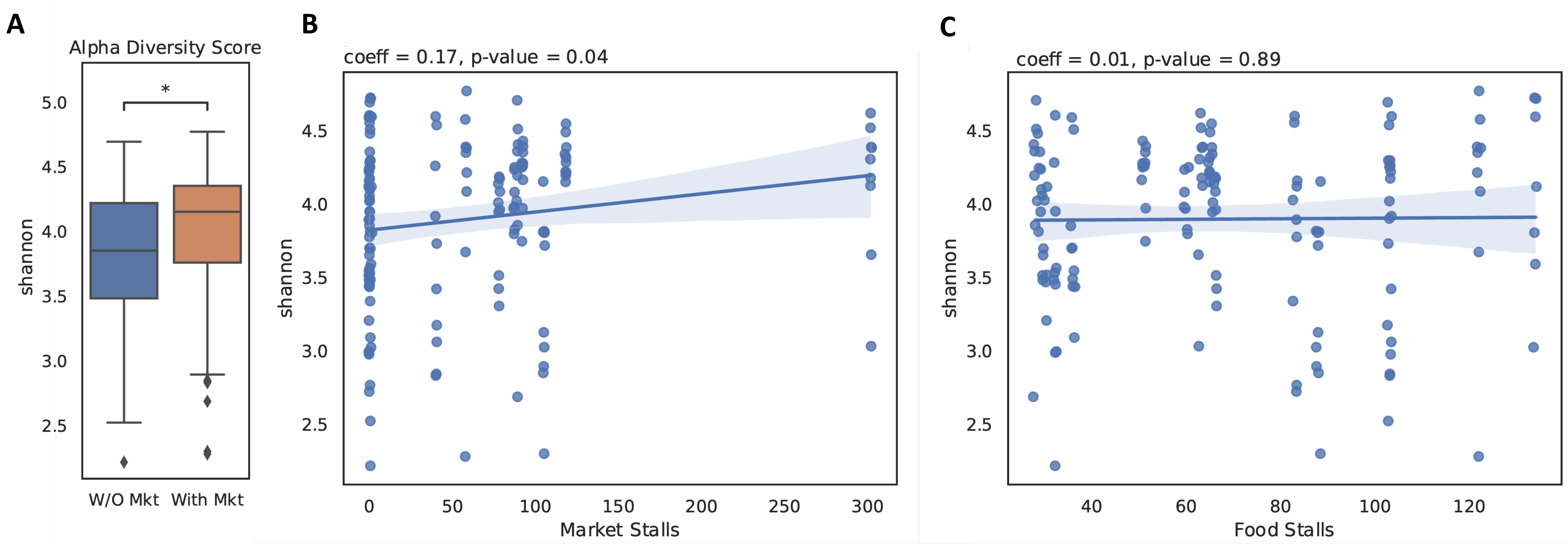


**Supplementary Figure 8:** Association of food centre features with microbial diversity. (A) Comparison of Shannon diversity of microbiomes in food centres with and without a wet market ("*": Wilcoxon test p-value<0.05). Linear regression plots showing relationship between Shannon diversity and number of (B) wet market stalls and (C) food stalls.

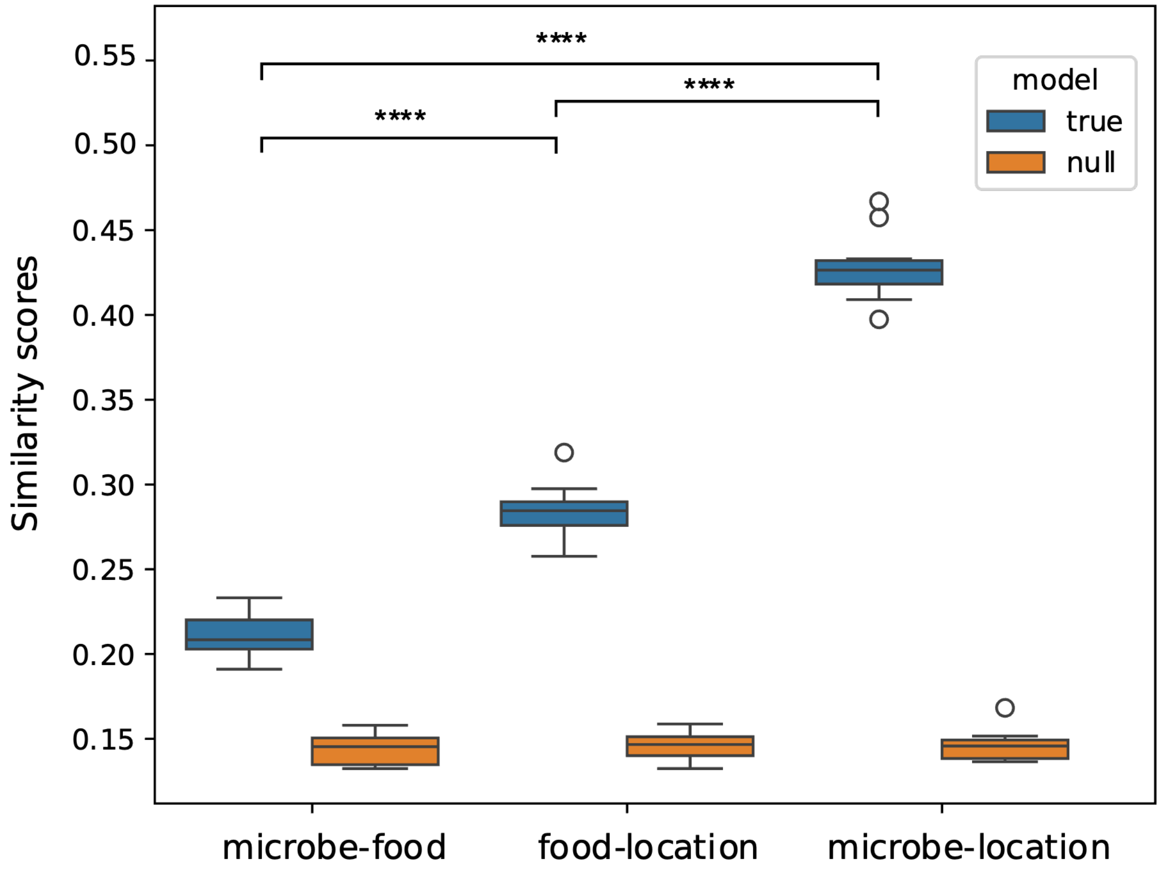


**Supplementary Figure 9:** Boxplots depicting similarity between clusters (true) obtained using microbial taxonomic profiles (Microbe), food-associated taxonomic profiles (Food), and sample locations (Location; **Methods**). We compared the similarity scores against a null model where the sample cluster labels are randomized (null). "****": Wilcoxon test p-value<0.0001.

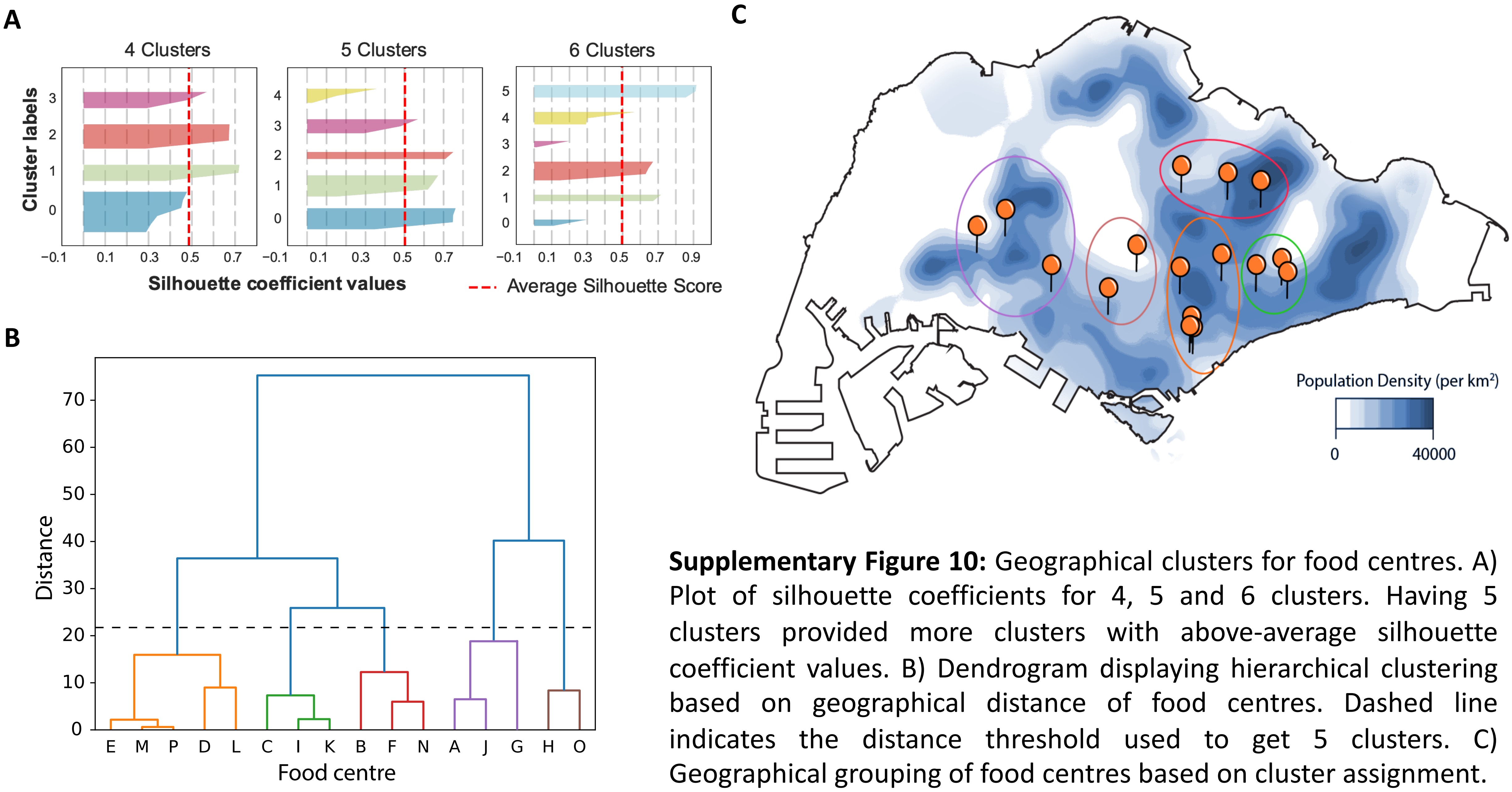


**Supplementary Figure 10:** Geographical clusters for food centres. A) Plot of silhouette coefficients for 4, 5 and 6 clusters. Having 5 clusters provided more clusters with above-average silhouette coefficient values. B) Dendrogram displaying hierarchical clustering based on geographical distance of food centres. Dashed line indicates the distance threshold used to get 5 clusters. C) Geographical grouping of food centres based on cluster assignment.

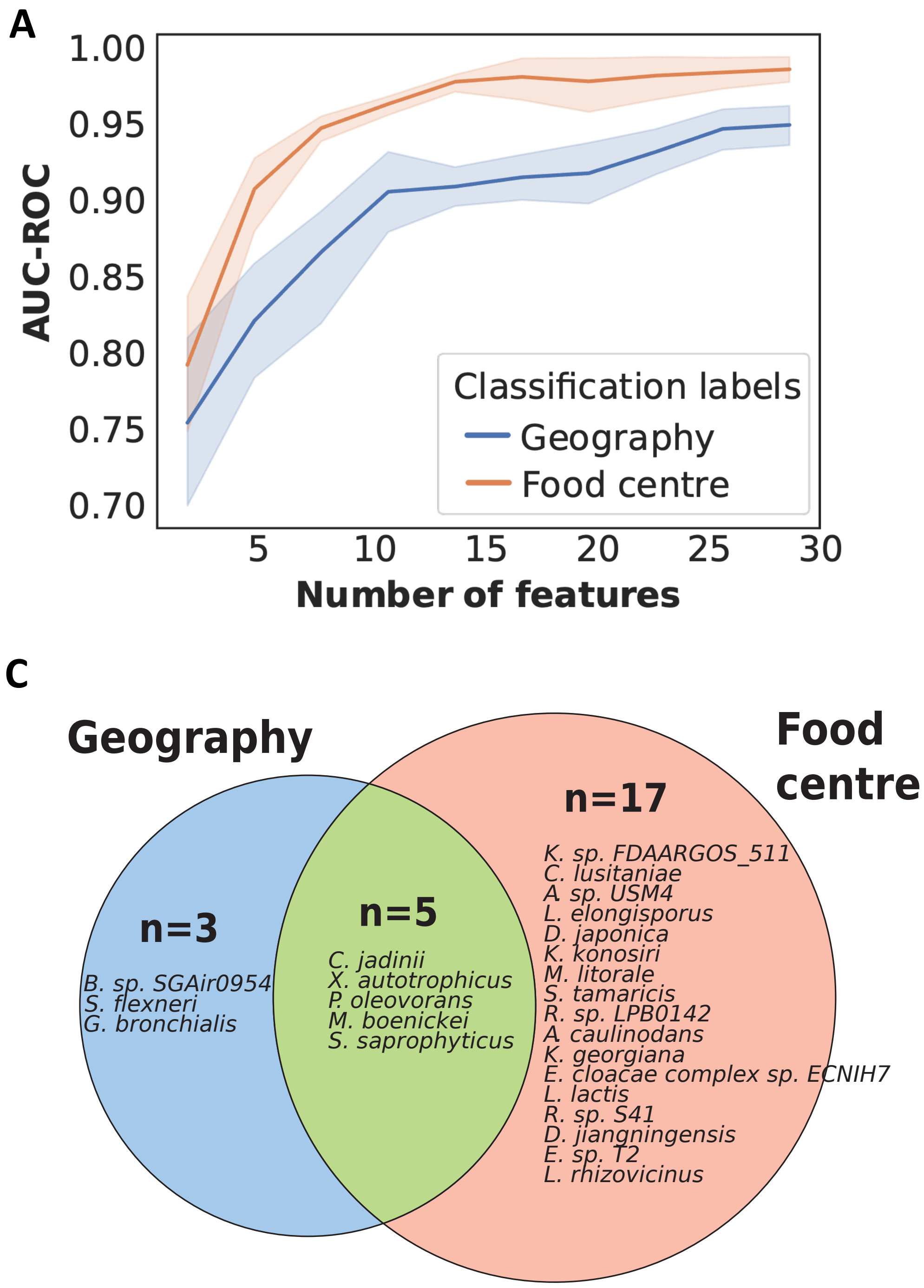


**Supplementary Figure 11:** Microbial signatures of geography. (A) Comparing AUC-ROC scores for classifying microbiome samples to their source of food centre location (orange) and geographical location (blue) with increasing number of features. (B) Relative abundance of microbes in the geographical signature. (C) Venn diagram comparing microbial features seen in geographical and food-court location signatures.

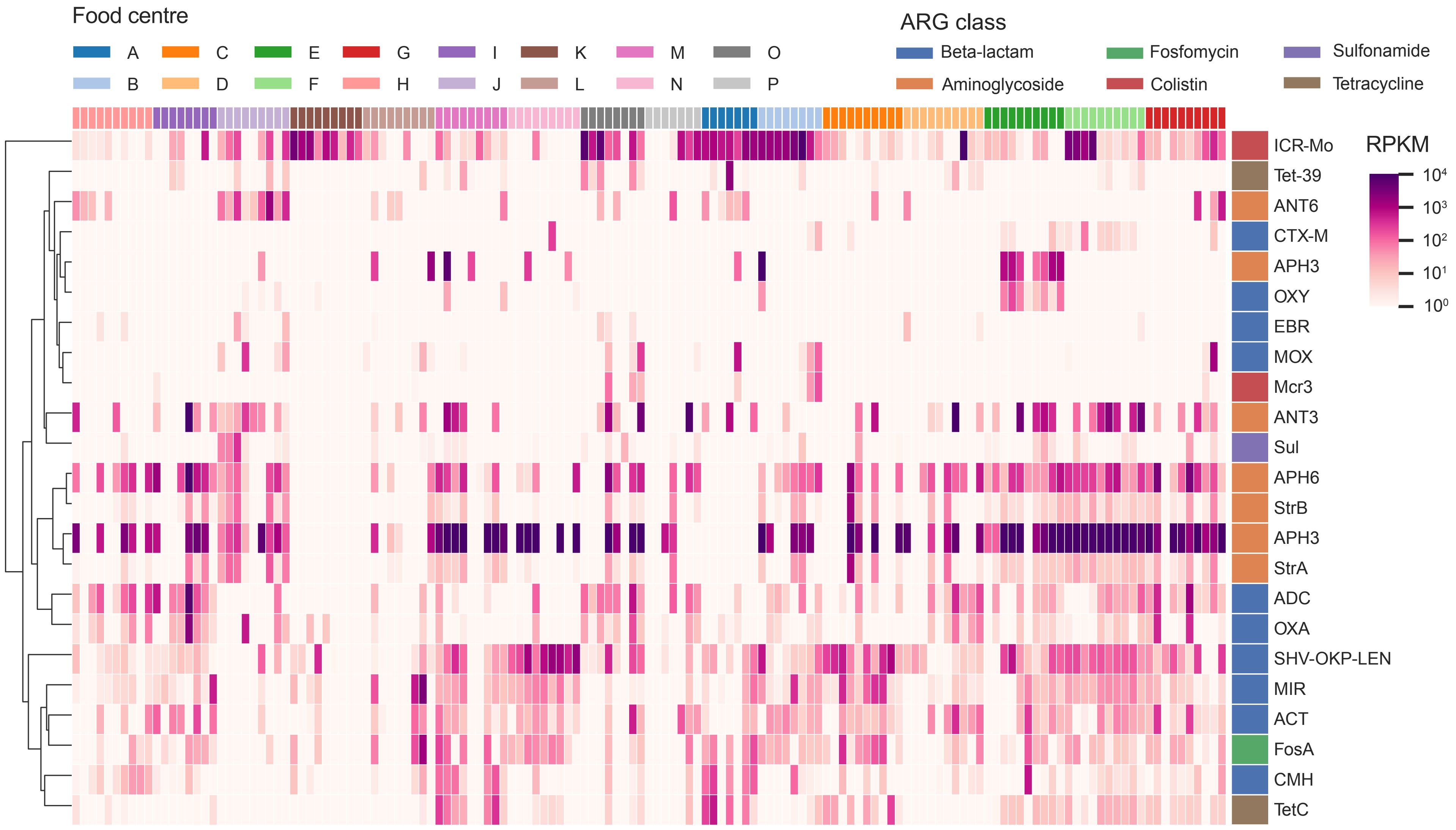


**Supplementary Figure 12:** Heatmap showing distribution of various antibiotic resistance gene classes (RPKM) in environmental metagenomes across sites in various food centres.

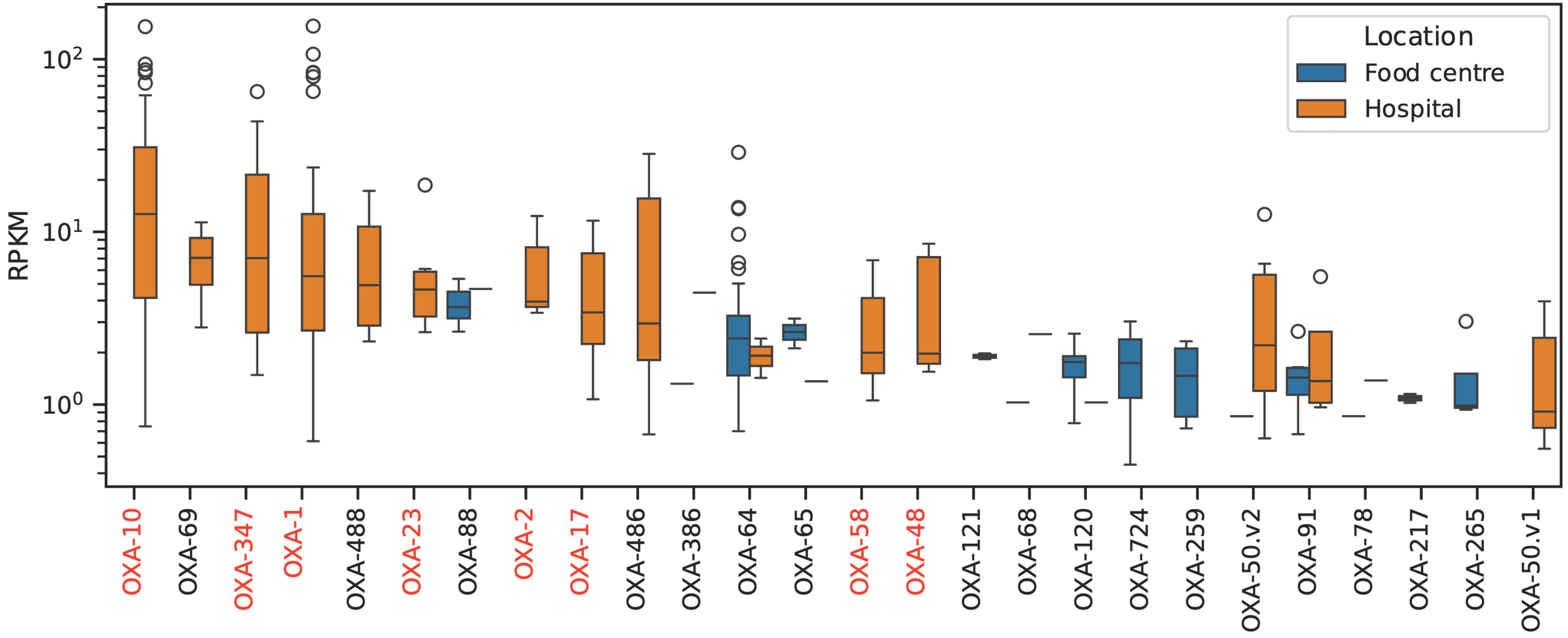


**Supplementary Figure 13:** Boxplots comparing the relative abundances (RPKM) of different OXA genes in food centre and hospital environment microbiomes. OXA genes that are associated with plasmids are highlighted in red.
